# Biguanides Associate with Decreased Early Mortality and Risk of Acute Kidney Injury In Hospitalized COVID-19 Patients: a nationwide retrospective cohort study in Japan

**DOI:** 10.1101/2024.07.20.24310736

**Authors:** Mari Sugimoto, Hiroaki Kikuchi, Eisei Sohara, Kavee Limbutara, Akihiro Hirakawa, Takayasu Mori, Koichiro Susa, Shuichiro Oya, Takefumi Suzuki, Soichiro Iimori, Kiyohide Fushimi, Shinichi Uchida

## Abstract

**Background:** Biguanide (BG) is the most-prescribed oral glucose-lowering medication worldwide and has potential for further therapeutic applications. The Coronavirus disease 2019 (COVID-19) pandemic is a global public health emergency. Nevertheless, there are still no established low-cost treatments against COVID-19, of which the morbidity and mortality rates varing from country to country. Therefore, a nationwide study of the COVID-19 affected population is essential to explore therapeutic effect of BG against COVID-19.

**Methods:** From the inpatient databases in Japan, covering the period from September 2021 to March 2023, which encompasses the era following the development of COVID-19 vaccines, we extracted data of 168,370 COVID-19 patients aged 20 to under 80 years who were suffered from diabetes mellitus treated with oral antidiabetic agents. The primary outcome was 100-day in-hospital mortality, and secondary outcome was the incidence of acute kidney injury (AKI) during hospitalization. We compared outcomes in patients who received BG with those in patients who did not, using a logistic regression analysis and Cox proportional hazards under both propensity score-unmatched and matched cohorts.

**Results:** The incidence of in-hospital death was significantly lower in the BG group (1.18 %) compared to the non-BG group (2.41%) (p < 0.001). Similarly, the incidence of AKI during hospitalization was significantly lower in the BG group (0.66 %) compared to the non-BG group (1.12%) (p < 0.001). Kaplan-Meier analysis from the propensity-score matched cohort showed a significantly better survival rate in the BG group (adjusted HR, 0.580; 95% CI 0.510-0.658; p < 0.001).

**Conclusion:** In COVID-19 patients, the use of oral biguanide use may be associated with a reduced in-hospital mortality and risk of AKI.

## Introduction

The novel coronavirus (2019-nCoV) is a newly emerging disease that was first reported in China and has subsequently spread worldwide (1). COVID-19 is caused by severe acute respiratory syndrome coronavirus 2 (SARS-CoV-2), which belongs to the Betacoronavirus genus (2). The challenge of COVID-19 is very high globally due to the complexity of its transmission and the lack of proven treatment. Although the current large-scale vaccine production has brought dawn to mankind, the globalization of vaccination (3) and virus mutation (4) are still challenging to eliminate SARS-CoV-2. While it is estimated that the overall case fatality ratio of the disease is 1.38% (95% confidence interval [Cl], 1.23–1.53), the absolute number of deaths have been remarkably high due to its high transmission rate and increased risk of death with age and comorbidities (5). While several treatments such as remdesivir and dexamethasone are either available or in development for severe COVID-19, interventions that can be administered early during the course of infection to prevent disease progression and longer-term complications are urgently needed.

Acute kidney injury (AKI) is a common complication in patients hospitalized for COVID-19 (6). AKI has been reported in more than one-fifth of COVID-19 hospitalizations, occurring twice as often in patients requiring intensive care unit treatment and being associated with approximately 10% dialysis requirement and high mortality (7).

Recent theoretical evidence has suggested that metformin, an oral biguanide, might have multiple beneficial effects, including anti-viral effect (8), improvement of the immune response (9), anti-cancer properties (10), and anti-hypertension effects (11). Metformin was discovered in 1922 (12) and introduced in the USA in 1995 and has emerged as the first choice and most-prescribed oral medication to lower blood glucose worldwide (12). Several clinical evidence have assessed the impact of metformin in COVID-19 patients, whereas their opinions were somehow discrepant (13)(14). This discrepancy may be due to the fact that prognosis of COVID-19 may be related to a complex combination of factors, such as infrastructure of medical facilities, resources of medical personnel, patient background (15), (16), (17). Therefore, to determine the factors which are associated with mortalities and to improve the management of COVID-19, a nationwide picture of the COVID-19 affected population is essential. DPC, which is an acronym for “Diagnosis Procedure Combination,” is a patient classification method developed in Japan for inpatients in the acute phase of illness of Japanese medical care, as well as evaluation and improvement of its quality (18).

Here, we carry out a retrospective analysis of the effect of in-hospital usage of biguanides on COVID-19 patients using nation-wide large patients’ cohort with 168,370 patients hospitalized with COVID-19. We considered the discharge or 100-day in-hospital death as the primary endpoint; incident of acute kidney injury as the secondary study endpoint, to gain an insight into biguanide treatment in COVID-19.

## Methods

### Study Design and Settings

This retrospective observational cohort study utilized data from nationwide administrative claims and a discharge abstract database in Japan, known as the DPC database. In brief, more than 1000 hospitals, including all 81 academic hospitals in Japan, contribute to the DPC database. The annual number of cases added to the database is nearly 7 000 000 and encompasses almost 50% of all hospital admissions in Japan (18). The DPC database contains the following information for each patient: demographics, a unique hospital identifier, diagnoses, outcomes, medications used, procedures performed, healthcare costs, and several disease-specific data. Diagnoses include the main diagnosis on admission, preexisting comorbidities, and postadmission complications, which are recorded separately with the International Classification of Diseases-10th (ICD-10) revision codes and text data in Japanese.

### Study participants

Patients were eligible for inclusion in the present study if they met the following criteria: (**1**) admitted to the hospitals participating in the DPC system from September 2021 to March 2023, **(2)** aged 20 to 80 years old, **(3)** had COVID-19 infection identified by the ICD-10 revision codes (Version 2013) of B-342, and **(4)** were treated with oral antidiabetic agents including biguanides, dipeptidyl peptidase-4 (DPP-4) inhibitors, sodium-glucose cotransporter type 2 (SGLT2) inhibitors, sulfonylureas (SU), alpha-glucosidase inhibitors (aGI), glucagon-like peptide-1 (GLP-1) analogs, thiazolidinediones (TZD).

Patients who reached the outcome within 2 days were excluded. The present analysis included 168,370 subjects (**Figure 1**). All patients’ data were followed up until they reached the outcome or until they were discharged from hospitals within 100 days after admission. For patients with multiple admissions at the same hospitals during the study period, only data from their first admission were used for subsequent analyses.

**Figure 1.**
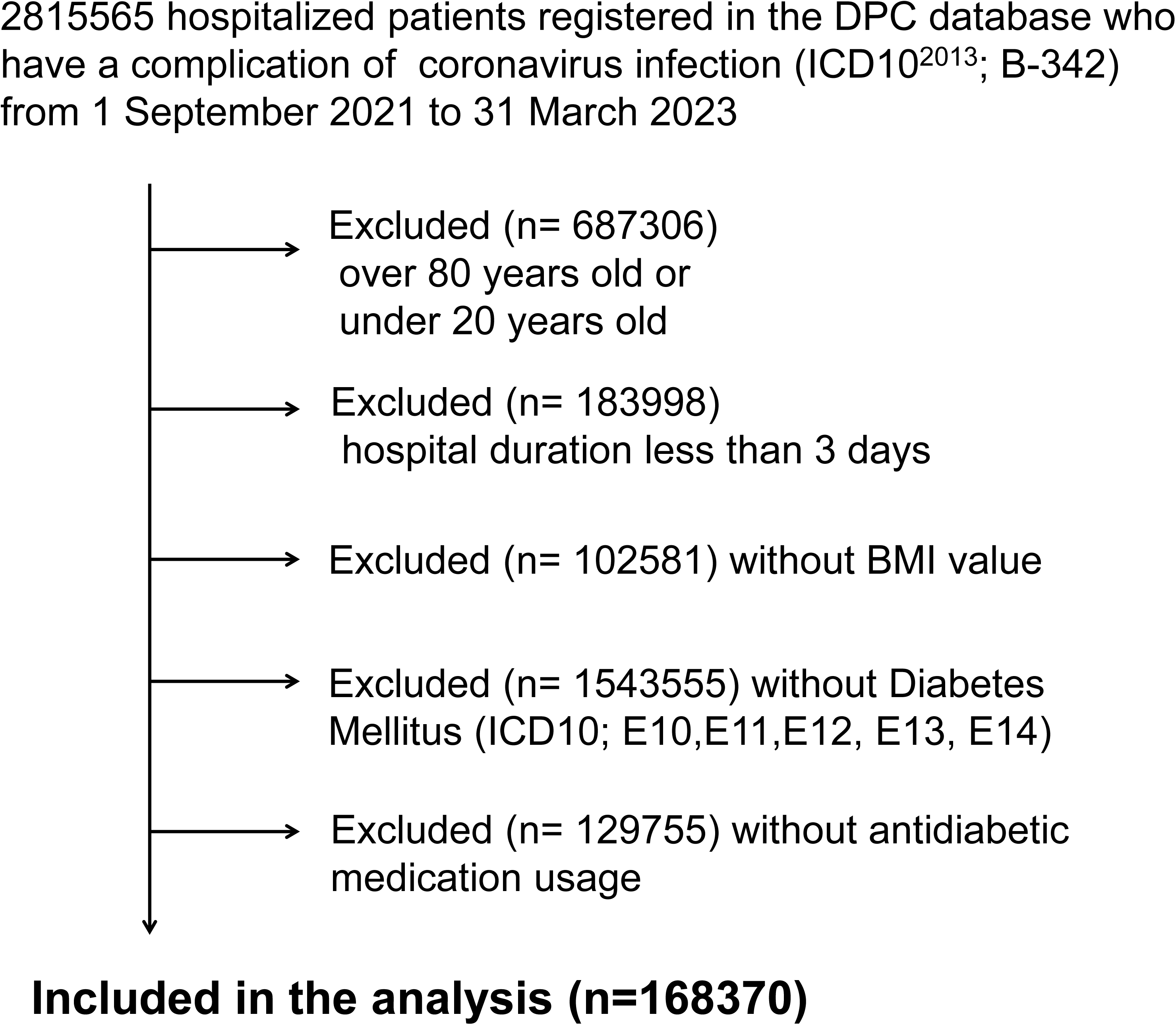
Flow chart of recruitment of participants and subjects studied. DPC, Diagnosis Procedure Combination; ICD, International Classification of Diseases; BMI, body mass index.

### Data Collection and Processing

Data on demographics, existing comorbidities, and several hospital characteristics were extracted for each patient based on pathophysiological importance. From the ICD-10 revision code (version 2013), existing comorbidities included hypertension, chronic kidney diseases, coronary artery disease (CAD), cerebral infarction (CI) and malignancies. Age, sex, body mass index (BMI) and Charlson score were also extracted from the DPC system. Medications, including BG, DPP-4 inhibitors, SGLT2 inhibitors, SU, aGI, GLP-1 analogs, TZD, and salicylate, were also included as hospital characteristics. All ICD-10 codes (**Supplementary Table S1**) and receipt numbers (**Supplementary Table S2**) for medication used in this study are summarized in Supplementary Tables.

### Study Groups and Study Endpoint

Patients were divided into two groups according to the presence (n = 30908) or absence (n = 137642) of biguanide treatment during hospitalization. The primary endpoint was the occurrence of in-hospital death. The secondary outcome was onset of acute kidney injury during hospitalization.

### Ethics

This study was approved by the Institutional Review Board of Tokyo Medical and Dental University (no. M2000-788) and conducted according to the principles of the Declaration of Helsinki. Informed consent was waived because of the anonymous nature of the data.

### Statistical analysis

Data acquisition and analysis were performed using R, version 4.0.2 (R Foundation for Statistical Computing). The values of normally distributed variables are presented as the mean ± standard deviation (SD). Categorical data are presented as counts and percentages. Intergroup comparisons were performed using the t-test or χ^2^ test, as appropriate. The primary analysis of interest was the relationship between BG usage and all-cause mortality. Multivariate logistic regression analysis was performed to identify variables that were independently associated with the primary outcome. The collinearities of candidate variables included in the multivariate logistic regression analysis were evaluated by the Pearson product-moment correlation coefficients (Pearson’s score). Kaplan–Meier survival curves were constructed, and log-rank testing was performed to assess the time to event of the primary outcome. A Cox proportional hazards model was used to evaluate the association between the BG groups and the primary outcome. The multivariate Cox proportional hazards models were adjusted for the demographics, comorbidities and medications for DM other than BG. The results are shown as hazard ratios (HRs) and 95% confidence intervals (CIs).

Propensity score matching was performed to achieve balance in covariates between patients treated with BG versus no BG therapy. Propensity scores were calculated using multivariable logistic regression models. Propensity scores included covariates that may affect both the likelihood of patients to receive the treatment of interest and the outcome of interest, and that were unbalanced between treatment groups before matching. Matching based on propensity scores was performed using a 1:1 nearest-neighbor algorithm, with a caliper width of 0.20. All statistical methods used are summarized in the **Supplementary Method**.

## Results

### Characteristics of the Cohort

A total of 168,370 patients hospitalized with COVID-19 infection who were treated with antidiabetic medication during 2021-2023 were selected for this study (**Figure 1**). The mean age of the cohort was 67.7 ± 10.6 years, 68.2% (*n* = 114,894) were males, and 8.79 % (*n* = 5,880) had chronic kidney disease (CKD). The mean BMI was 24.8 ± 5.44 kg/m^2^. The baseline characteristics of the patients between two groups, namely, “Patients without biguanide (BG) treatment (No Biguanide)” and “Patients with Biguanide (Biguanide)” were presented in **Table 1**. Of the 168370 patients, 30908 (18.4 %) received BG. Patients with BG treatment were younger, more likely to be females, had higher Charlson score, higher BMI, a higher prevalence of cerebral infarction (CI); and a lower prevalence of hypertension, CKD, cardiovascular disease, malignancies, and pneumonia (**Table 1**). Patients with BG treatment was more likely to be treated with SU and TZD, and less likely treated with DPP4 inhibitors, SGLT2 inhibitors and salicylate (**Table 1**)

**Table 1.**
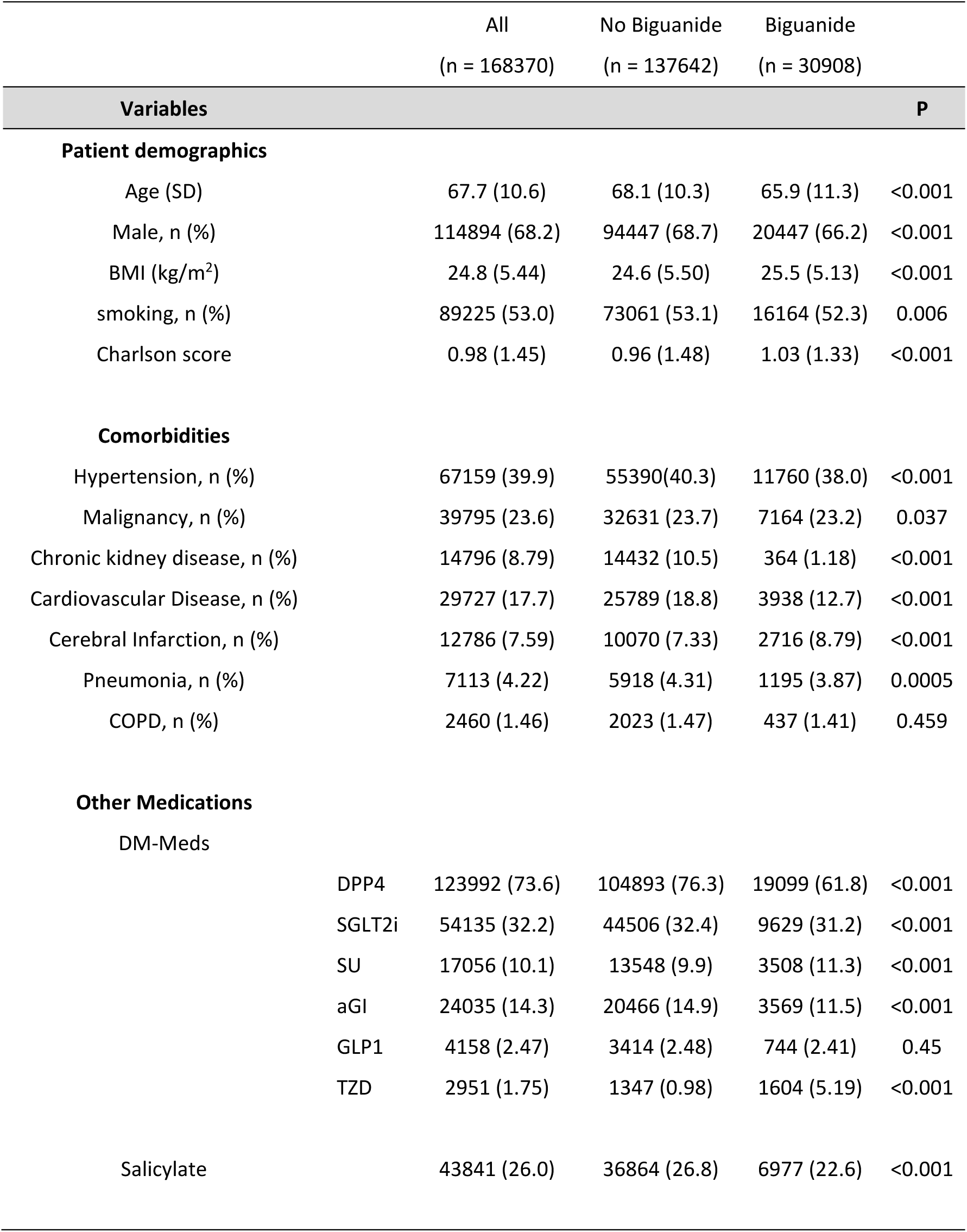
Comparison of baseline population characteristics between groups.

The values of normally distributed variables are presented as the mean ± standard deviation (SD). Categorical data are presented as counts and percentages. CAD, coronary artery disease; CKD, chronic kidney disease; BMI, body mass index; COPD, chronic obstructive pulmonary disease; DM, diabetes mellitus; DPP4, Dipeptidyl peptidase-4 inhibitors; SGLT2i, sodium-glucose cotransporter type 2 inhibitors; SU, sulfonylureas; aGI, alpha-glucosidase inhibitors; GLP-1, glucagon-like peptide-1 analogs, TZD, thiazolidinediones.

### In-hospital BG usage is associated with lower in-hospital mortality rates in admitted COVID-19 patients under unmatched cohort

Among the 168,370 patients included in the analysis, the primary endpoint of in-hospital death occurred in 3689 patients (2.19%) during the 100 days follow-up period. In the univariate, unadjusted analysis, patients who received BG were less likely to experience the primary endpoint event compared to those who did not (odds ratio, 0.49; 95% CI, 0.44 to 0.54) (**Table 2**). Next, we sought to identify variables significantly associated with the primary outcome. All Pearson’s scores were less than 0.5, therefore, there is no strong correlation between any two variables listed in **Table 1**. In multivariate logistic regression analyses, BG usage was significantly associated with lower mortality in all models. (**Table 2**).

**Table 2.**
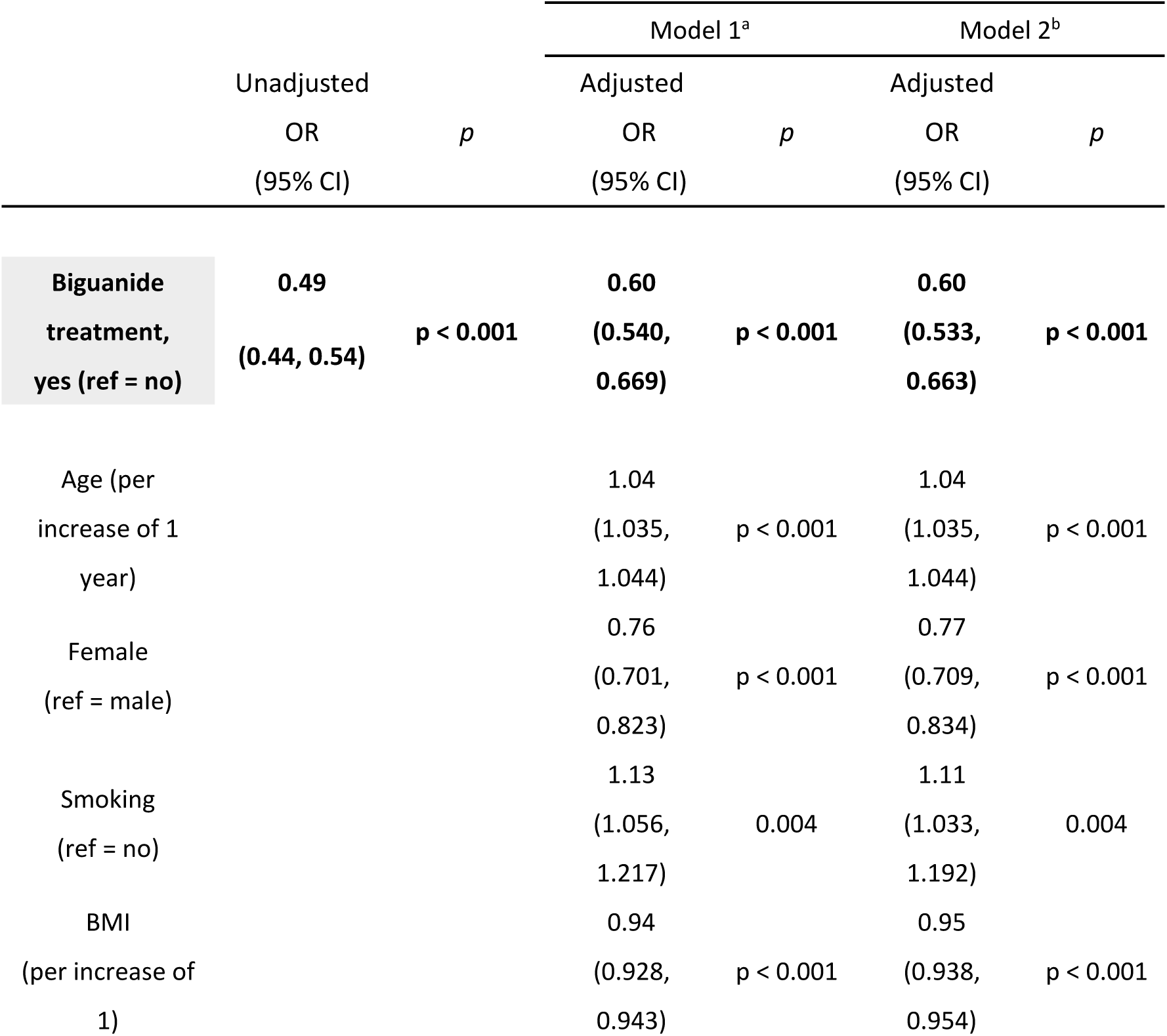
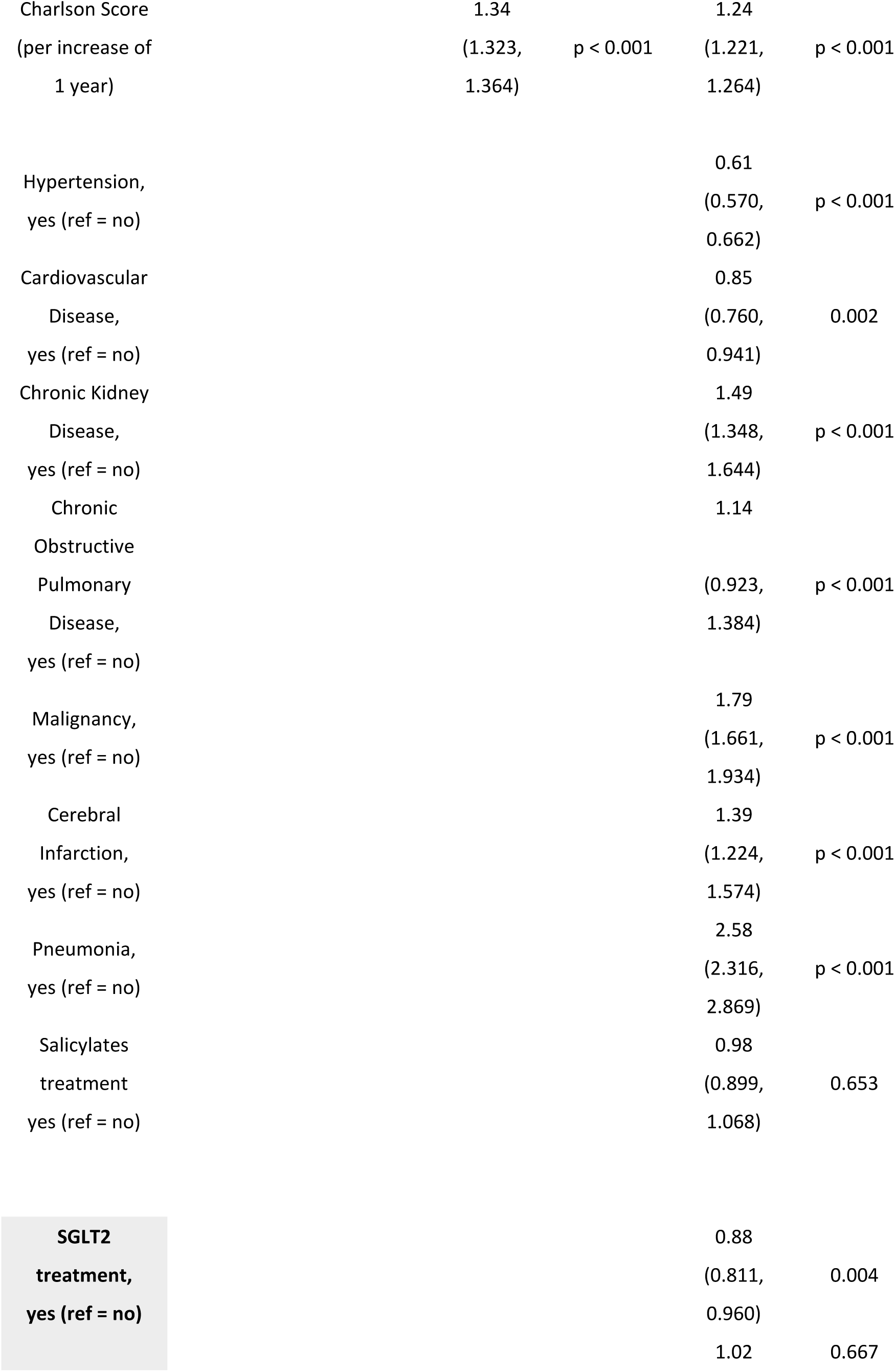
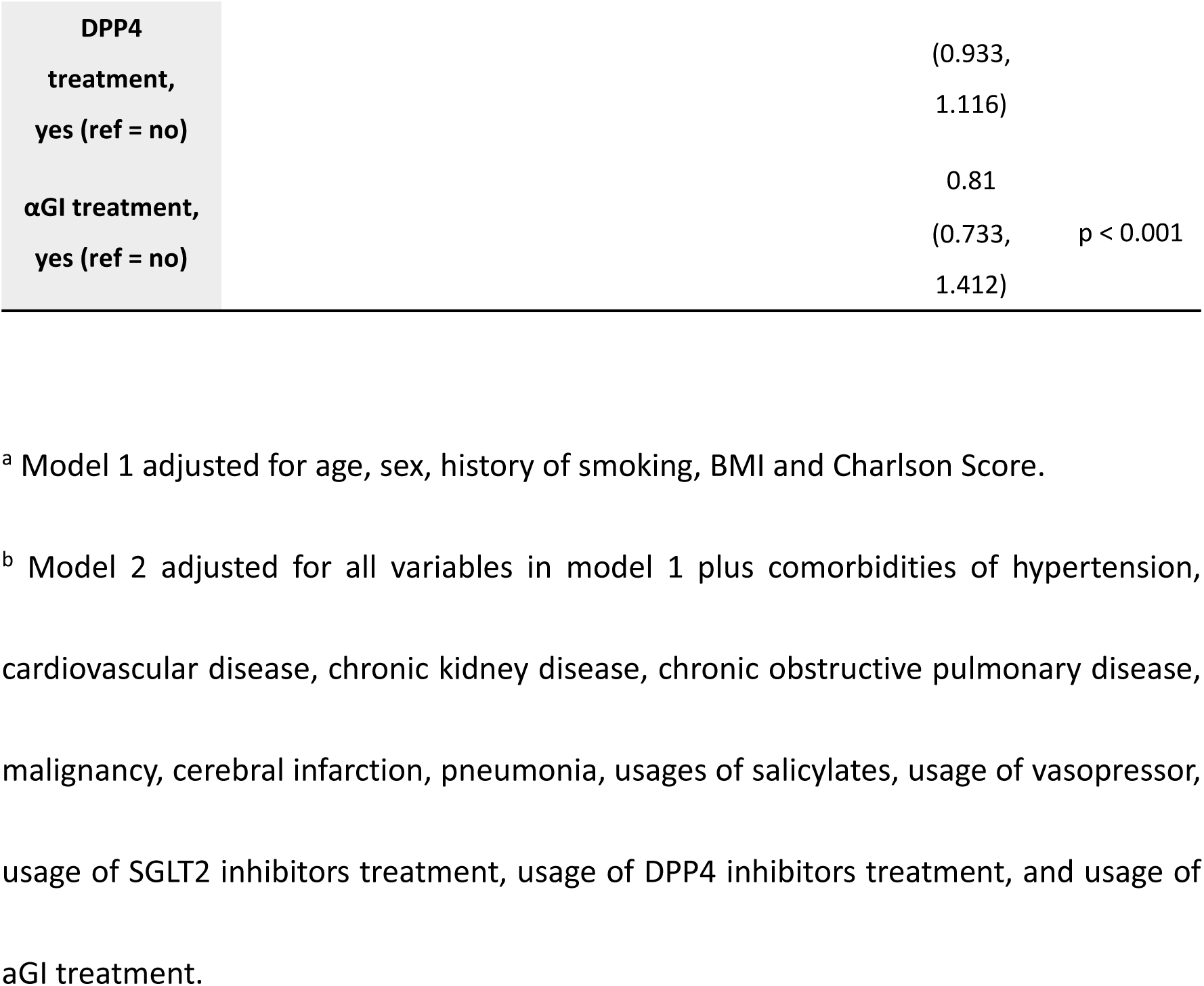
Association of BG usage and the primary outcome of in-hospital death in the univariate analysis and multivariable analysis in the overall study cohort.

|  | Unadjusted |  | Model 1<sup>a</sup> |  | Model 2<sup>b</sup> |  |
| --- | --- | --- | --- | --- | --- | --- |
|  | OR<br>(95% CI) | *p* | Adjusted<br>OR<br>(95% CI) | *p* | Adjusted<br>OR<br>(95% CI) | *p* |
| **Biguanide**<br>**treatment,**<br>**yes (ref = no)** | **0.49**<br>**(0.44, 0.54)** | **p < 0.001** | **0.60**<br>**(0.540, 0.669)** | **p < 0.001** | **0.60**<br>**(0.533, 0.663)** | **p < 0.001** |
| Age (per<br>increase of 1<br>year) |  |  | 1.04<br>(1.035,<br>1.044) | p < 0.001 | 1.04<br>(1.035,<br>1.044) | p < 0.001 |
| Female<br>(ref = male) |  |  | 0.76<br>(0.701,<br>0.823) | p < 0.001 | 0.77<br>(0.709,<br>0.834) | p < 0.001 |
| Smoking<br>(ref = no) |  |  | 1.13<br>(1.056,<br>1.217) | 0.004 | 1.11<br>(1.033,<br>1.192) | 0.004 |
| BMI<br>(per increase of<br>1) |  |  | 0.94<br>(0.928,<br>0.943) | p < 0.001 | 0.95<br>(0.938,<br>0.954) | p < 0.001 |

|  |  |  |  |  |
| --- | --- | --- | --- | --- |
| Charlson Score<br>(per increase of<br>1 year) | 1.34<br>(1.323,<br>1.364) | p < 0.001 | 1.24<br>(1.221,<br>1.264) | p < 0.001 |
| Hypertension,<br>yes (ref = no) |  |  | 0.61<br>(0.570,<br>0.662) | p < 0.001 |
| Cardiovascular<br>Disease,<br>yes (ref = no) |  |  | 0.85<br>(0.760,<br>0.941) | 0.002 |
| Chronic Kidney<br>Disease,<br>yes (ref = no) |  |  | 1.49<br>(1.348,<br>1.644) | p < 0.001 |
| Chronic<br>Obstructive<br>Pulmonary<br>Disease,<br>yes (ref = no) |  |  | 1.14<br>(0.923,<br>1.384) | p < 0.001 |
| Malignancy,<br>yes (ref = no) |  |  | 1.79<br>(1.661,<br>1.934) | p < 0.001 |
| Cerebral<br>Infarction,<br>yes (ref = no) |  |  | 1.39<br>(1.224,<br>1.574) | p < 0.001 |
| Pneumonia,<br>yes (ref = no) |  |  | 2.58<br>(2.316,<br>2.869) | p < 0.001 |
| Salicylates<br>treatment<br>yes (ref = no) |  |  | 0.98<br>(0.899,<br>1.068) | 0.653 |
| **SGLT2**<br>**treatment,**<br>**yes (ref = no)** |  |  | 0.88<br>(0.811,<br>0.960) | 0.004 |
|  |  |  | 1.02 | 0.667 |

|  |  |  |
| --- | --- | --- |
| DPP4 treatment, yes (ref = no) | (0.933, 1.116) |  |
| αGI treatment, yes (ref = no) | 0.81 (0.733, 1.412) | p < 0.001 |

<sup>a</sup> Model 1 adjusted for age, sex, history of smoking, BMI and Charlson Score.
<sup>b</sup> Model 2 adjusted for all variables in model 1 plus comorbidities of hypertension, cardiovascular disease, chronic kidney disease, chronic obstructive pulmonary disease, malignancy, cerebral infarction, pneumonia, usages of salicylates, usage of vasopressor, usage of SGLT2 inhibitors treatment, usage of DPP4 inhibitors treatment, and usage of αGI treatment.

In a Kaplan–Meier analysis, COVID-19 patients with BG treatment had a significantly higher survival rate compared to those without BG treatment (*P* < 0.001, Log-rank chi-square = 155) (**Figure 2**). Cox proportional-hazards model analysis also showed that BG therapy was associated with a reduction in mortality (HR, 0.574; 95% CI 0.516-0.639; p < 0.001) (**Figure 3**).

**Figure 2.**
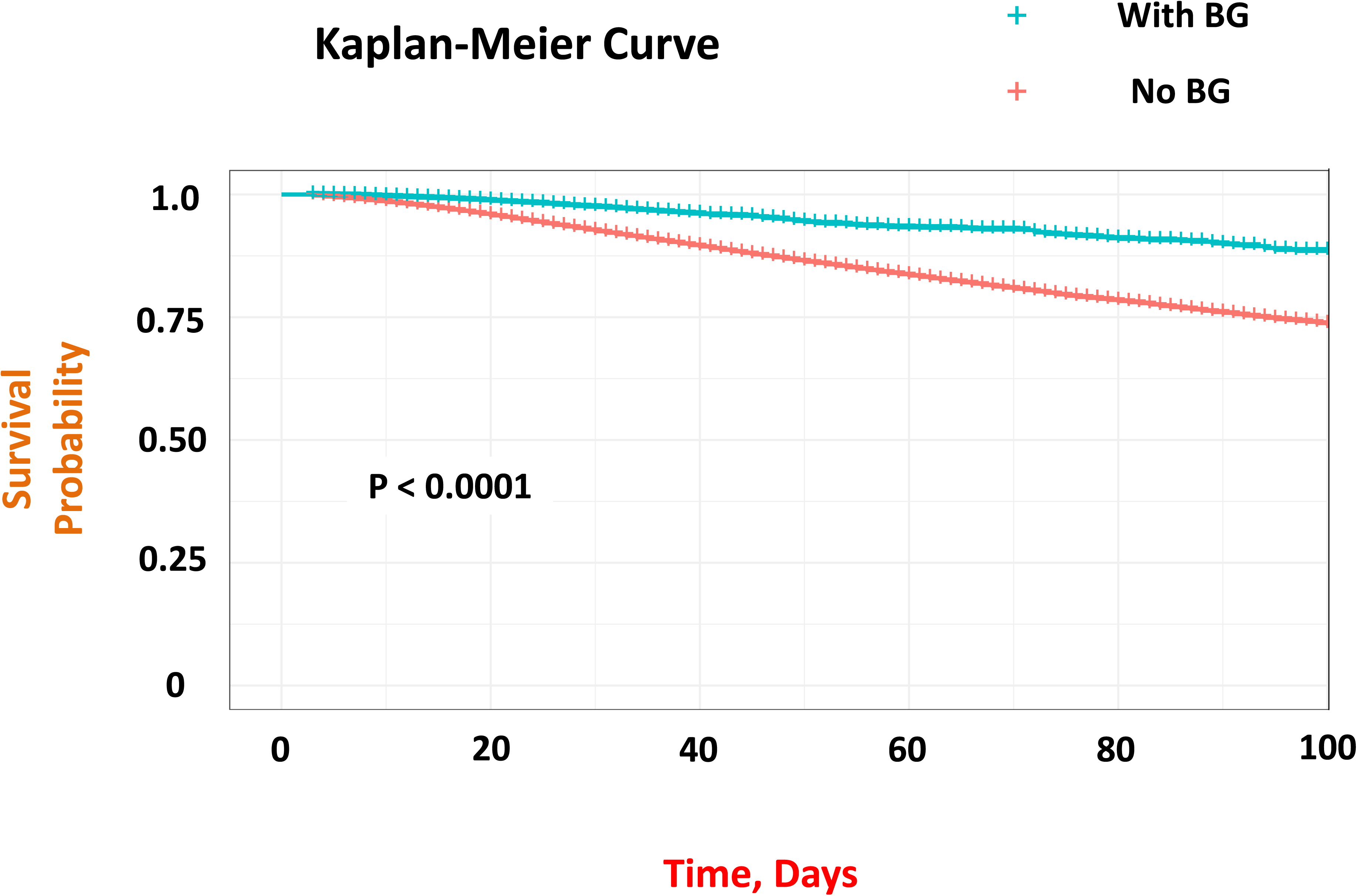
Kaplan–Meier estimates of all-cause mortality for admitted COVID patients with biguanide usage and without biguanide usage under unmatched cohort. COVID-19 patients with BG treatment had a statistically significant higher survival rate than COVID-19 patients without BG treatment (*p*< 0.001, Log-rank chi-square = 155) BG: biguanide.

**Figure 3.**
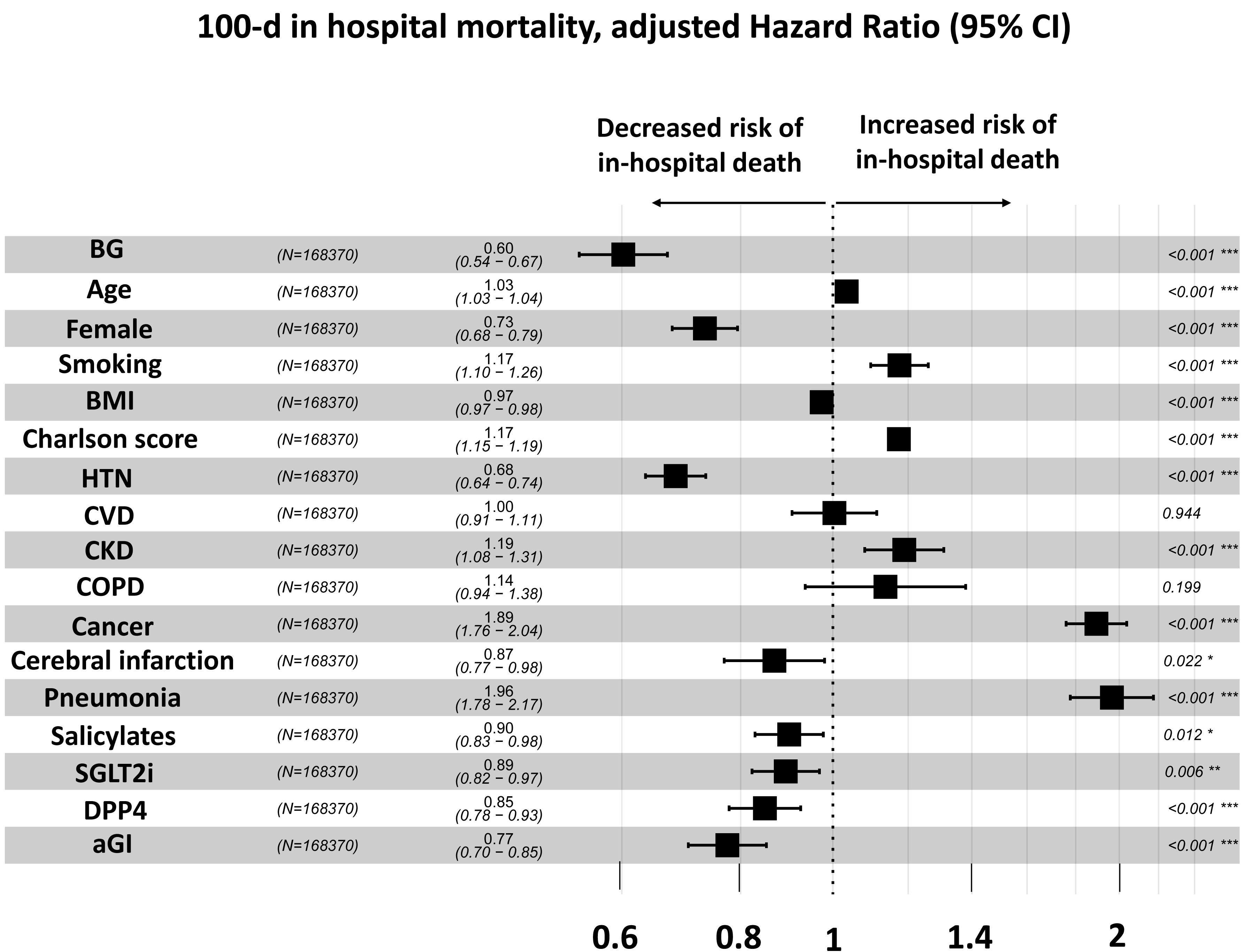
A forest plot showing the hazard ratio and 95% confidence intervals associated with variables with time to the primary endpoint (100d in-hospital death) Black squares represent the hazard ratios, and the horizontal bars extend from the lower limit to the upper limit of the 95 % confidence interval of the estimate of the hazard ratio. BG, biguanide; BMI, body mass index; HTN, hypertension; CVD, cardiovascular disease; CKD, chronic kidney disease; COPD, chronic obstructive pulmonary disease; SGLT2i, sodium-glucose cotransporter type 2 inhibitors (SGLT2); DPP4, Dipeptidyl peptidase-4 inhibitors; aGI, alpha-glucosidase inhibitors.

### In-hospital BG usage is associated with lower incidence rate of AKI in admitted COVID-19 patients under unmatched cohort

Among the 168,370 patients included in the analysis, the secondary endpoint of onset of AKI developed in 1746 (1.04 %). In the univariate, unadjusted analysis, patients who received BG were less likely to experience the secondary endpoint event compared to those who did not (odds ratio, 0.59; 95% CI, 0.504 to 0.676) (**Table 3**). In multivariate logistic regression analyses, BG usage was significantly associated with a lower rate of onset of AKI in all models. (**Table 3**).

**Table 3.**
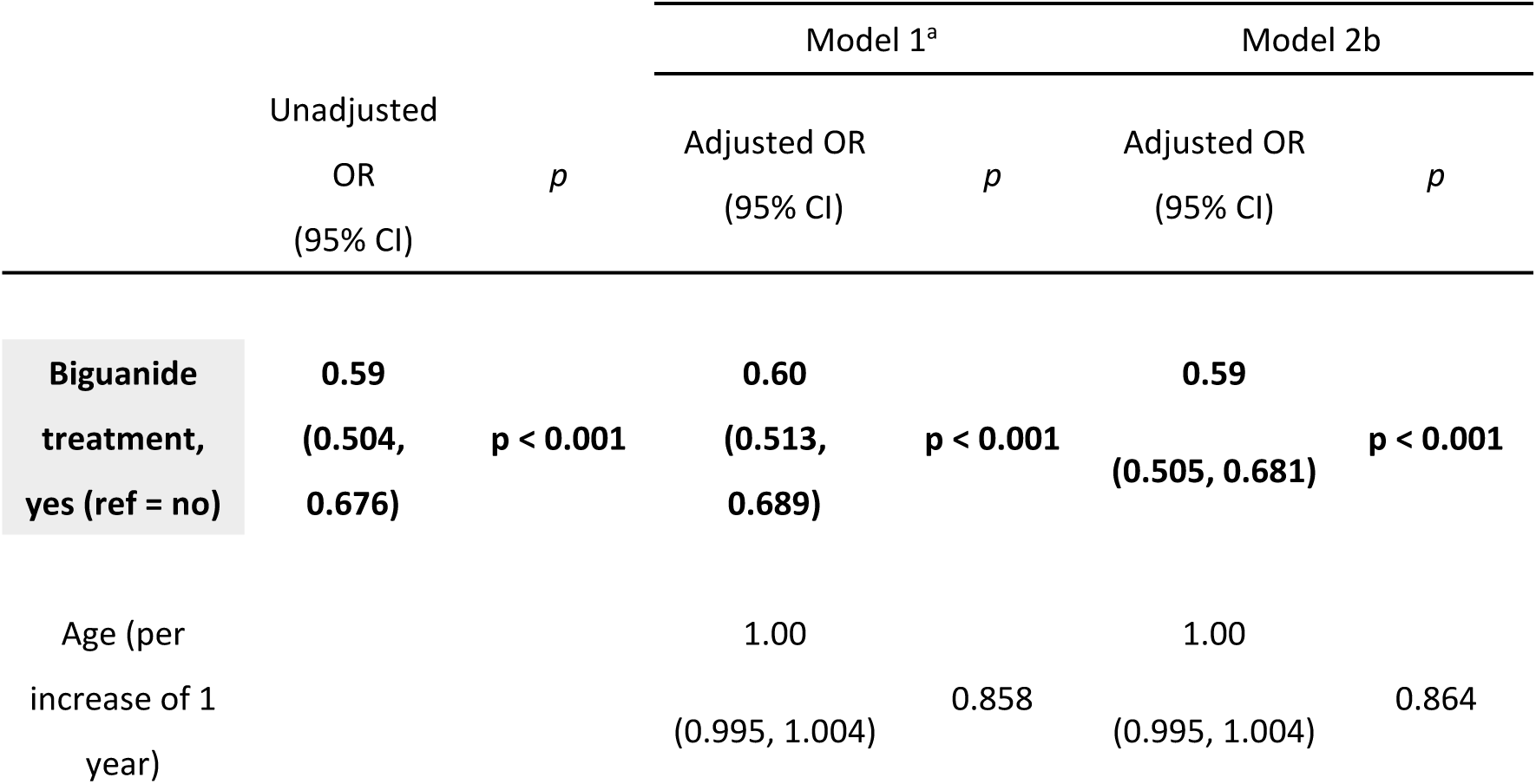
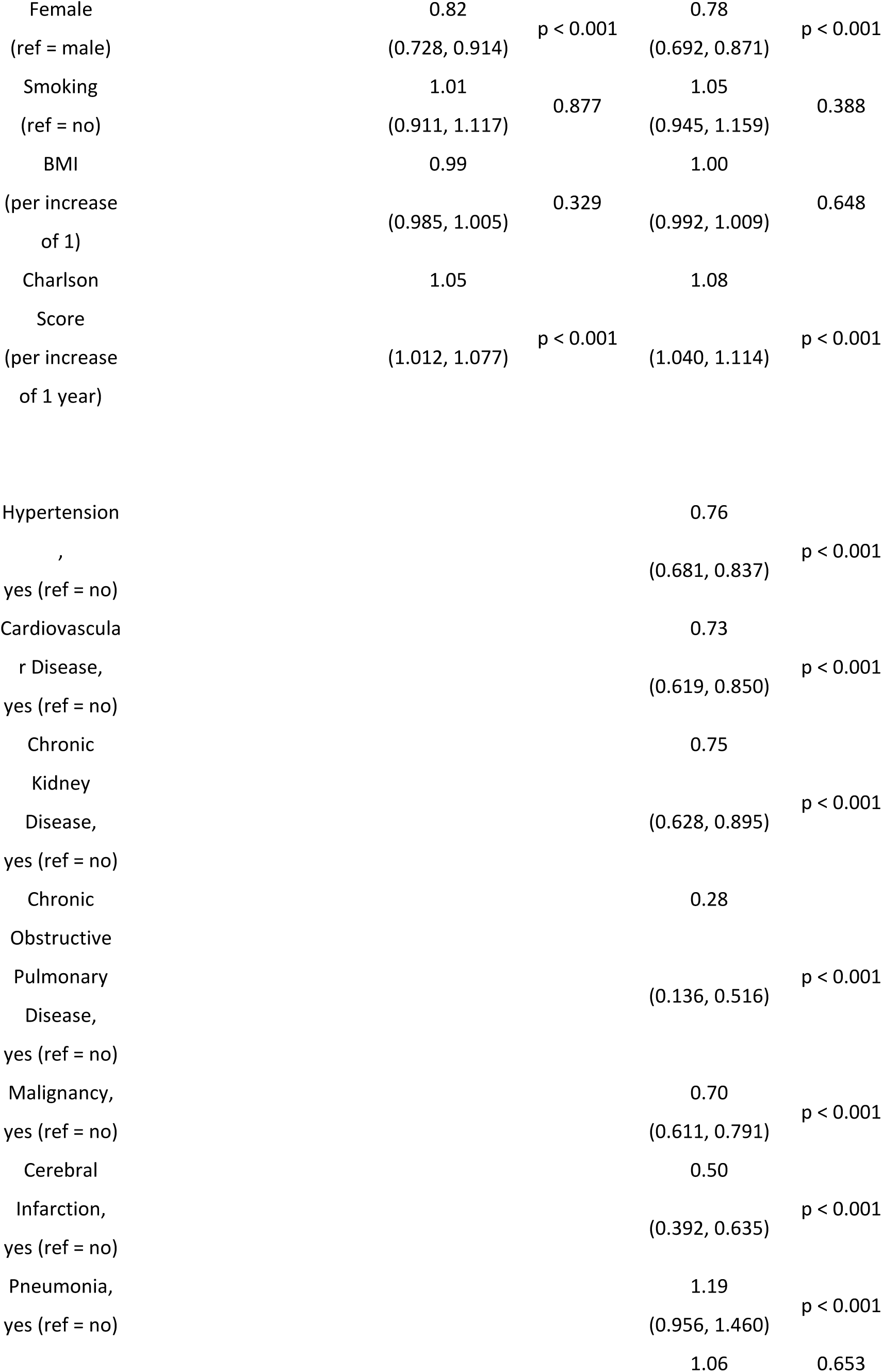
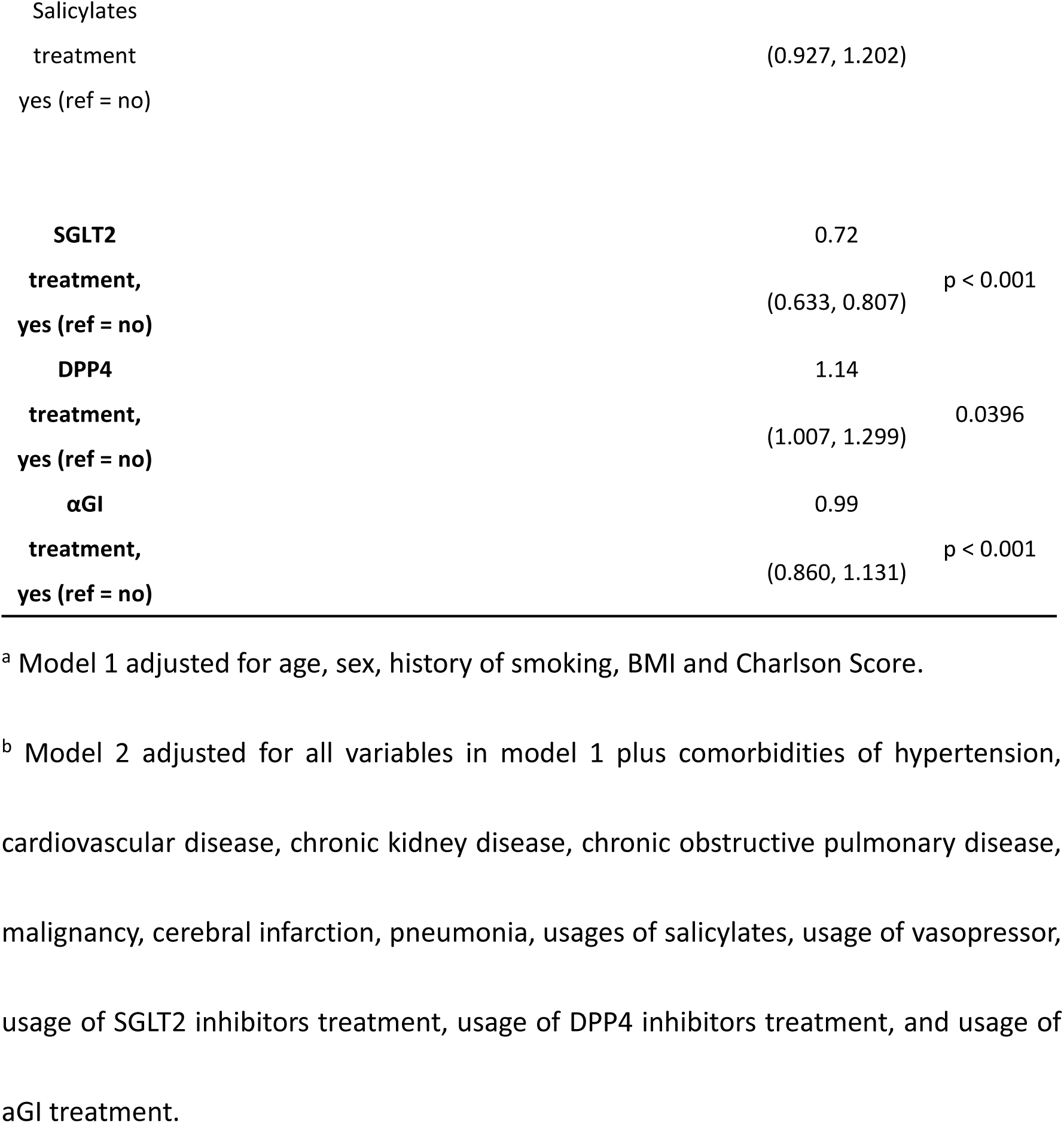
Association of BG usage and AKI-onset in the univariate analysis and multivariable analysis in the overall study cohort.

### In-hospital BG usage is associated with lower in-hospital mortality rates and with lower incident rate of AKI in admitted COVID-19 patients under propensity-score matched cohort

A major limitation in retrospective studies is the bias in the likelihood of patients receiving the treatments being studied. In unadjusted observational studies, disease severity is a confounding factor affecting treatment decisions and outcomes, often precluding accurate analysis of potential treatment effects. To address this, propensity score matching for disease severity and other variables has been utilized in some observational studies, leading to findings compatible with those obtained from randomized controlled trials (19), (20), (21). In the clinical setting, some patients may not receive BG due to the risk of lactic acidosis, particularly with metformin, which is called metformin-associated lactic acidosis (MALA) (22). Though MALA is an extremely rare condition, MALA can be induced by several conditions that either increase the production of lactate or decrease its clearance. These conditions include renal impairment, liver disease, heart failure and malignancy. Tumors can produce large amounts of lactate through anaerobic glycolysis, known as Warburg effect (23) Thus, we utilized propensity score matching and multivariable regression analysis incorporating markers of disease severity and other clinical covariates including age, sex, BMI, Charlson score, comorbidities of malignancy, chronic kidney disease, cardiovascular disease, pneumonia, COPD and use of medications (i.e., DPP4 inhibitors, SGLT2 inhibitors and aGI) (**See also Supplementary method**). Accordingly, 30908 patients were allocated to the group exposed to BG and 30908 were not exposed. The differences between BG and clinical covariables were attenuated in the propensity-score-matched samples as compared to the unmatched samples. (**Table 4**, **Supplementary Figure 1**).

**Table 4.** Characteristics of patients receiving or not receiving BG, after propensity-score matching.

| Propensity<br>matched data | All<br>(n = 61816) | No Biguanide<br>(n = 30908) | Biguanide<br>(n = 30908) |  |
| --- | --- | --- | --- | --- |
| Variables |  |  |  | Absolute<br>Standardized<br>Difference |
| **Patient demographics** |  |  |  |  |
| Age (SD) | 65.2 (11.9) | 64.5 (12.4) | 65.9 (11.3) | 0.06 |
| Male, n (%) | 40915 (66.2) | 20468 (66.2) | 20447 (66.2) | 0.02 |
| BMI (kg/m<sup>2</sup>) | 25.7 (5.74) | 25.9 (6.28) | 25.5 (5.13) | 0.04 |
| smoking, n (%) | 32343 (52.3) | 16179 (52.3) | 16164 (52.3) | - |
| Charlson score | 0.80 (1.35) | 0.85 (1.38) | 0.75 (1.31) | 0.03 |
| **Comorbidities** |  |  |  |  |
| Hypertension<br>(%) | 23505 (38.0) | 11745 (38.0) | 11760 (38.0) | - |
| Malignancy,<br>n (%) | 13776 (22.3) | 6612 (21.4) | 7164 (23.2) | 0.01 |
| Chronic kidney<br>disease, n (%) | 741 (1.20) | 377 (1.22) | 364 (1.18) | 0.01 |
| Cardiovascular Disease, n (%) | 7598 (12.3) | 3660 (11.8) | 3938 (12.7) | 0.04 |
| CI event, n (%) | 4970 (8.04) | 2254 (7.29) | 2716 (8.79) | - |
| Pneumonia, n (%) | 2451 (3.96) | 1256 (4.06) | 1195 (3.87) | 0.01 |
| COPD, n (%) | 790 (1.28) | 353 (1.14) | 437 (1.41) | 0.01 |
| **Other Medications** |  |  |  |  |
| DM-Meds |  |  |  |  |
| DPP4 | 38303 (61.9) | 19204 (62.1) | 19099 (61.8) | 0.03 |
| SGLT2 | 21834 (35.3) | 12205 (39.5) | 9629 (31.2) | 0.12 |
| SU | 7759 (12.6) | 4251 (13.8) | 3508 (11.3) | - |
| aGI | 7674 (12.4) | 4105 (13.3) | 3569 (11.5) | 0.07 |
| GLP1 | 2486 (4.02) | 1742 (5.64) | 744 (2.41) | - |
| TZD | 2142 (3.47) | 538 (1.74) | 1604 (5.19) | - |
| Salicylate, n (%) | 13753 (22.2) | 6776 (21.9) | 6977 (22.6) | - |

Next, using this propensity score-matched group of 61816 patients, we explored the effects of in-hospital BG usage. From logistic regression analysis, patients who had received BG were less likely to have had a primary endpoint event than those who did not (odds ratio, 0.62; 95% CI, 0.548 to 0.708). Cumulative incidence curves also showed a significant reduction in in-hospital death among propensity score-matched patients who were treated with BG (**Figure 4**). Cluster-paired Cox proportional-hazards regression model analysis also showed that BG therapy was associated with reduction in mortality (**hazard ratio, 0.580; 95% CI 0.510-0.658; p < 0.001**). Regarding the secondary endpoint of onset of AKI, logistic regression analysis showed that patients who had received BG were less likely to have had a secondary endpoint event than those who did not (**odds ratio, 0.64; 95% CI, 0.538** - **0.767**).

**Figure 4.**
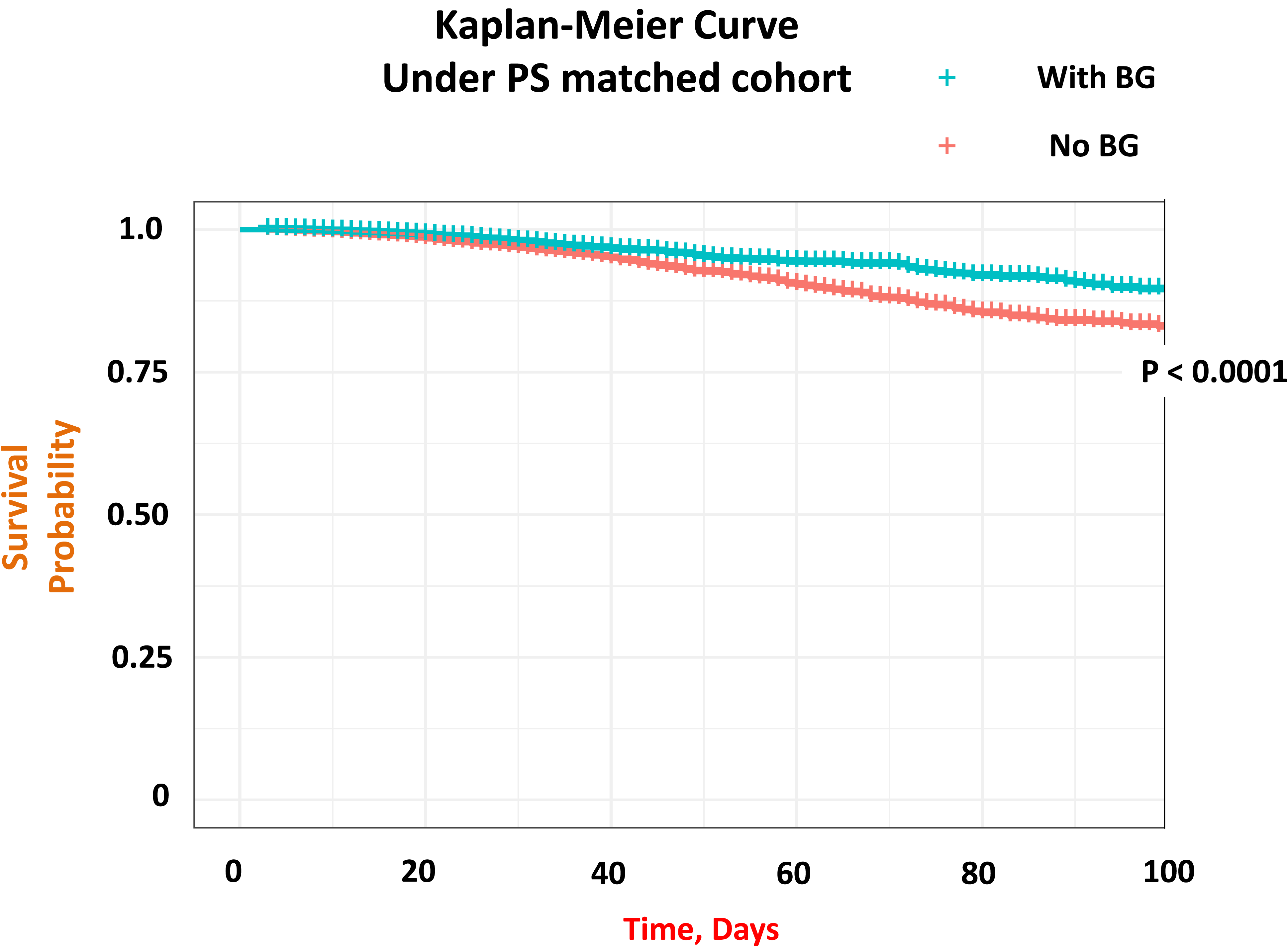
Kaplan–Meier estimates of all-cause mortality for admitted COVID patients with biguanide usage and without biguanide usage under propensity-score matched cohort. COVID-19 patients with BG treatment had a statistically significant higher survival rate than COVID-19 patients without BG treatment (*p* < 0.001, Log-rank chi-square = 64.5) even in the propensity-score matched cohort. BG: biguanide.

## Discussion

In our nationwide study of hospitalized DM-COVID-19 patients in Japan, we demonstrated that in-hospital biguanide usage is significantly associated with decreased mortality and a lower incident rate of AKI. In addition to the multivariable regression analysis for unmatched cohort, we utilized propensity score matching and multivariable regression analysis to diminish treatment selection bias by generating treatment and control groups with well-balanced covariates. This allowed for a more reliable comparison of potential biguanide treatment effects. Our findings suggest a beneficial role for biguanide in the treatment of hospitalized COVID-19 patients. To the best of our knowledge, this is the first observational study to suggest the association between biguanide usage and decreased rate of AKI development in the hospitalized COVID-19 patients.

The emergence of SARS-CoV-2 created a worldwide public health emergency. SARS-CoV-2, the causative agent of COVID-19, was first isolated during an outbreak in Wuhan, China, in December 2019 (1). Due to its rapid dissemination to several countries and virtually all continents, the World Health Organization (WHO) declared a pandemic in March 2020. COVID-19 has caused almost 800 million cases of disease worldwide since January 2020, and over 7 million deaths (https://covid19.who.int). This is 100,000 times as many cases as SARS in 2003, and 10,000 times as many deaths. Currently, about 30% of the world population, including 70% of people in low-income countries, remain unvaccinated against COVID-19 (24). Thus, although the WHO ended their public health emergency for COVID-19 on May 11, 2023, COVID-19 remains a pandemic as of July 2024 (25).

Although several antiviral agents, including Nirmatrelvir/ritonavir, molnupiravir, azvudine and remdesivir are approved as COVID-19 therapeutics (https://www.fda.gov/drugs/emergency-preparedness-drugs/coronavirus-covid-19-drugs), we still face challenges in terms of global accessibility, especially in developing countries, due to regulatory hurdles, economic limitations, and other barriers. Therefore, it remains highly desirable to find new COVID-19 treatments that are not only more efficient and safer but also more accessible and affordable to help combat and ultimately end the pandemic. Drug repurposing is an essential strategy, as this approach can save a considerable amount of time and money since the pharmacokinetics, pharmacodynamics and safety profiles of these drugs are already established, ((26), (27)).

The biguanide metformin is the most prescribed drug for type 2 diabetes (T2D) taken by an estimated 150 million individuals worldwide. Several studies identified metformin as a potential therapeutic agent because of its possible action against proteins involved in the mRNA translation process, its antiviral activity in vitro, and its anti-inflammatory and antithrombotic activities ((28), (29)). However, the therapeutic effect of metformin for COVID-19 remains controversial. Although several observational studies have suggested that patients who were receiving metformin as treatment for diabetes at the time of their COVID-19 diagnosis had a lower risk of progressing to severe COVID-19 ((30), (31)), a few other clinical trials (TOGETHER trial, COVID-OUT trial) implied that the use of metformin did not reduce the risk of hospitalization or death ((32), (33)). Thus, although we demonstrated the association of in-hospital usage of biguanide with a lower rate of in-hospital death using nationwide large cohort database, the therapeutic effects of metformin for COVID-19 patients need further scrutiny.

Among the pleiotropic effects of metformin beyond its hypoglycemic effect, reno-protective effect through the activation of 5-adenosine monophosphate-activated protein kinase (AMPK) in renal tubular epithelium has recently attracted attention ((34), (35), (36)). AMPK activity is exquisitely sensitive to cellular energy stress, which is reflected in the rising AMP level and an increase in the AMP–adenosine triphosphate (ATP) ratio, indicating that AMPK is a critical cellular energy sensor (37). As the kidneys consume a large amount of energy to regulate body fluids and blood pressure and to excrete waste products and toxins, dysregulation of AMPK leads to impairment of the renal tubular homeostasis(38). Indeed, we recently reported that dysregulation of AMPK signal in the proximal tubule induced tubular atrophy and fibrosis in the subtotal nephrectomy mouse model ((39), (40)). AMPK has also been reported to have the direct effect on virus entry via viral receptor angiotensin-converting enzyme 2 (ACE2) (41). AMPK modifies ACE2 by phosphorylating ACE2 at Ser680 in human umbilical vein endothelial cells and human embryonic kidney 293 (HEK293T) cells(42). Several kidney-specific RNA-Seq datasets provide detailed information about ACE2 gene expression in kidney cell types. The data indicates that ACE2 mRNA is strongly expressed in proximal tubule epithelial cells ((43), (44)) but not in more distal renal tubule cells, podocytes, mesangial cells or glomerular endothelial cells (45). Thus, it could be proposed that AMPK induces a conformational change in the ACE2 receptor in the proximal tubular cells, leading to decreased binding affinity with the SARS-CoV-2, hence reduced infectivity and a reduced risk of severe disease (46). The mechanisms underlying the reno-protective effects of metformin for COVID-19 induced renal damage should be clarified in the future.

In summary, this is the first study showing that in-hospital BG usage affects both in-hospital mortality and the incident rate of AKI in COVID-19 patients in Japan. These findings provide a foundation for future studies to clarify the mechanisms of metformin’s reno-protective role in COVID-19 patients.

## Data Availability

All data produced in the present study are available upon reasonable request to the authors

## Acknowledgments

This work was supported by Grant-in-Aid for Research Activity Start-up (23K19610) to H. Kikuchi, Grant-in-Aid for Early-Career Scientists (24K19143) to H. Kikuchi, Japan Agency for Medical Research and Development (AMED), Grant Number 24ek0310023h0001 to H. Kikuchi, and the Uehara Memorial Foundation to H. Kikuchi.

## Author contributions

M.S. and H.K. contributed equally to this work, and either has the right to list her/himself first in bibliographic documents. M.S. and H.K designed the research. M.S., H.K., K.L., A.H, S.O. and K.F. performed experiments and statistical analysis. M.S., H.K., K.L., A.H., E.S., T.M., K.S., S.O, S.I. and S.U. participated in the discussions and interpretation of the data. M.S., H.K., and S.O. wrote the manuscript.

## List of supplementary material

- Supplementary methods

- Supplementary Figure S1

- Supplementary Table S1

- Supplementary Table S2

