## Supplementary Information for "Biguanides Associate with Decreased Early Mortality and Risk of Acute Kidney Injury In Hospitalized COVID-19 Patients: a nationwide retrospective cohort study in Japan"

### Supplementary method:

#### Data extraction from DPC system

For the present study we used the Japanese DPC Database(1), which is a patient discharge and administrative claims data base to which the acute care hospitals under the DPC reimbursement policy voluntarily submit. The DPC database is similar to the US Medicare claims database (2) or the US Nationwide Impatient Samples (3). The DPC has been conducted by the DPC Research Group funded by the Ministry of Health, Labour and Welfare, Japan. As of April 2020, the payment system initially introduced to 82 hospitals in 2003 has been applied to 1,757 hospitals with a total of 483,180 beds. This number is thought to be enough to cover almost all acute inpatients and is about 30% of all hospitals with beds for general patients (including those in subacute care and rehabilitation, but excluding those with mental illness, infectious disease, tuberculosis, and long-term care) and about 54% of all beds of hospitals with beds for general patients across the country (1). The DPC database can be used to identify, track, and analyze national trends in health-care utilization, access, quality, outcomes, and costs. The database includes the following information: location of hospital; patient age and sex; diagnoses, comorbidities at admission and complications after admission coded with International Classification of Diseases, 10th Revision codes; procedures coded according to the Japanese claim classification (K-codes); drugs and devices used; length of stay (LOS); in-hospital mortality; and costs. The diagnoses are recorded by the physicians in charge with reference to the medical charts.

We selected all the DPC hospitals that participated in the DPC survey every year between 2021 and 2023 which cover the era following the development of COVID-19 vaccines. In Japan, vaccination for COVID-19 was introduced in February 2021. All patients who were diagnosed with COVID-19 infection (DPC 2013 code B-342) were enrolled in the present study.

**Statistical methods used**

Comparison of baseline population characteristics with continuous variables (**Table 1**) including age, BMI, Charlson score are analyze by unpaired- T test using t.test () function in R. Comparison of baseline population characteristics with categorical variables (**Table 1**) including Sex, Smoking, Hypertension, Malignancy, Chronic kidney disease, Cardiovascular disease, Cerebral infarction, Pneumonia, COPD

DM-meds (DPP4, SGLT2, SU, aGI, GLP1, TZD) , Salicylate were analyzed by chi-squared test using chisq.test () function in R. Association of BG usage and the primary outcome and secondary outcome (**Table 2,3**) were analyzed by logistic regression analysis using glm () function in R. Kaplan–Meier analysis was performed using survfit () function. Logrank test was performed using Surv () function in R. Cox proportional hazard model analysis was performed using coxph () function in R. Propensity Score Matching (**Table 4**) was performed using a 1:1 nearest-neighbor algorithm, with a caliper width of 0.20 using matchit () function in R.

### Supplementary Figure

#### Supplementary Figure 1

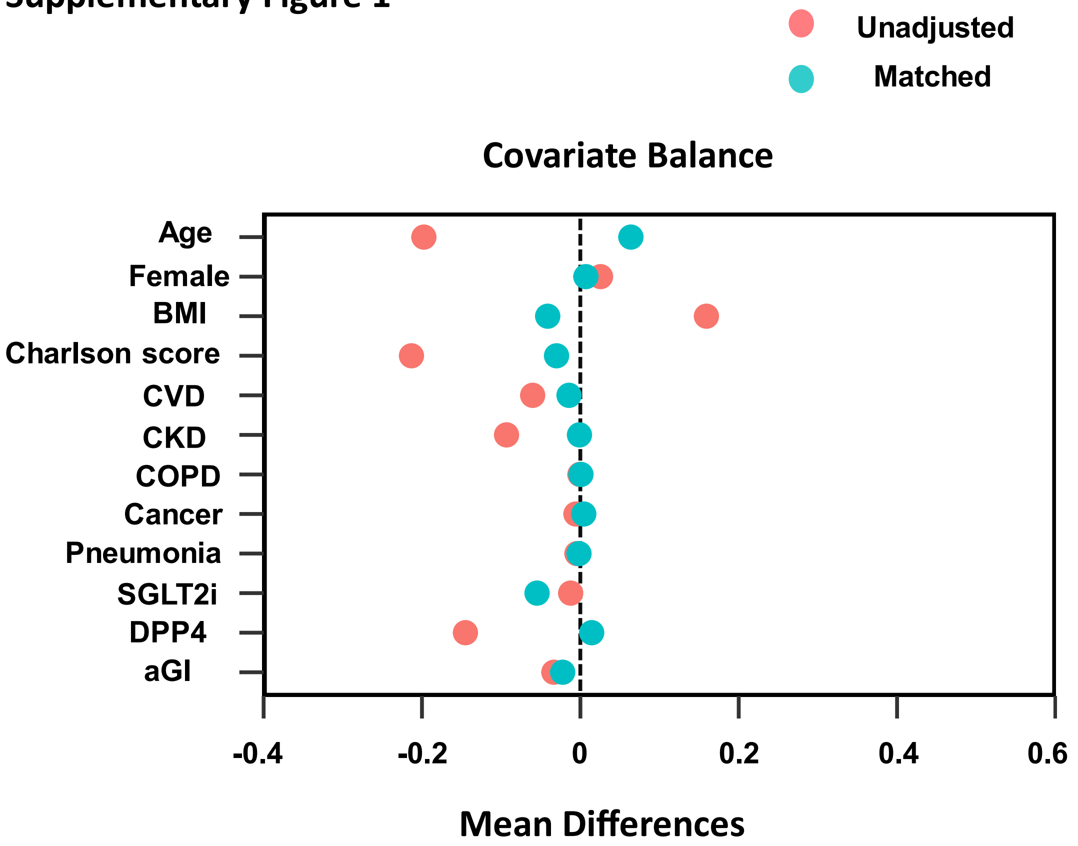


**Supplementary Figure 1. Standardized mean differences in the unmatched and matched sample**

### Supplementary Table

#### Supplementary Table 1. ICD-10 codes used for this study

| *Diseases* | ICD-10 codes (2013 version) | | | | | | | | |
|---|---|---|---|---|---|---|---|---|---|
| **Chronic kidney disease** | N18.0 | N18.8 | N18.9 | N19 | | | | | |
| **Acute kidney injury** | N17.0 | N17.1 | N17.2 | N17.8 | N17.9 | | | | |
| **Diabetes Mellitus** | E10.X | E11.X | E12.X | E13.X | E14.X | | | | |
| **Hypertension** | I10 | I11.X | I12.X | I13.X | I15.X | | | | |
| **Cardiovascular Disease** | I20.X | I21.X | I22.X | I23.X | I24.X | I25.X | | | |
| **Solid Malignancy** | C00.X | C01 | C02.X | C03.X | C04.X | C05.X | C06.X | C07.X | C08.X | C09.X |
| | C10.X | C11.X | C12 | C13.X | C14.X | C15.X | C16.X | C17.X | C18.X | C19 |
| | C20 | C21.X | C22.X | C23 | C24.X | C25.X | C26.X | | | |
| | C30.X | C31.X | C32.X | C33 | C34.X | C37 | C38.X | C39.X | | |
| | C40.X | C41.X | C43.X | C44.X | C45.X | C46.X | C47.X | C48.X | C49.X | |
| | C50.X | C51.X | C52 | C53.X | C54.X | C55 | C56 | C57.X | C58 | |
| | C60.X | C61 | C62.X | C63.X | C64.X | C65 | C66 | C67.X | C68.X | C69.X |
| | C70.X | C71.X | C72.X | C73.X | C74.X | C75.X | | | | |
| | D00.X | D01.X | D02.X | D03.X | D04.X | D05.X | D06.X | D07.X | D09.X | |
| **Non-solid malignancy** | C81.X | C82.X | C83.X | C84.X | C85.X | C88.X | C89 | | | |
| | C90.X | C91.X | C92.X | C93.X | C94.X | C95.X | C96.X | | | |
| **Cerebral Infarction** | I63.X | I66.X | | | | | | | | |
| **Pneumonia** | J15.X | J18.X | J84.X | | | | | | | |
| **Chronic obstructive pulmonary disease** | J44.X | | | | | | | | | |
| **COVID-19 infection** | **B34.2** | | | | | | | | | |

### Supplementary Table 2. Receipt numbers used for this study

| *Treatment* | Receipt numbers for treatment | | | | | | | | | |
|---|---|---|---|---|---|---|---|---|---|---|
| **Biguanide (BG)** | 621974701 | 621974701 | 621974701 | 622242501 | 622242501 | 622242501 | 622517101 | 622450401 | 622450401 | 622450301 |
| | 622450301 | 621986301 | 621986301 | 621986301 | 620004480 | 620004480 | 620004480 | 610444147 | 610444147 | 610444147 |
| | 620004502 | 620004502 | 620004502 | 613960014 | 613960014 | 613960014 | 613960015 | 613960015 | 613960015 | |
| **Salicylate** | 620000485 | 620000486 | 611140017 | 611140019 | 620000483 | 611140794 | 620001952 | 610431009 | 620002629 | 610441006 |
| | 620000484 | 611140849 | 611140795 | 610421011 | 620000487 | 620000488 | 611140798 | 620004280 | 610408009 | 611140793 |
| | 611140850 | 611140849 | 810000000 | | | | | | | |
| | 620000065 | 611140181 | 616220021 | 620001612 | 620001562 | 662650007 | 662650002 | 662650015 | 662650092 | 662660006 |
| | 662640125 | 662650002 | 662650001 | 662650001 | 660441023 | 662650108 | 660421082 | 662650093 | 660408074 | 660408076 |
| | 641140027 | 641140028 | 641140079 | 641140113 | 620004727 | 610407146 | 610463004 | 620004897 | 616210018 | 620008624 |
| | 620008997 | 666210001 | 622023001 | 622173901 | 621969902 | 610409338 | 620008070 | 620506801 | 621867201 | 622234801 |
| | 660470002 | 622442701 | 622517001 | 620008510 | 610443053 | 622341601 | 610443056 | 610443057 | 621375001 | 621362001 |
| **Thiazolidinediones (TZD)** | 610432040 | 610432041 | 621990901 | 621991001 | 621986301 | 621986301 | 622048401 | 622048501 | 622086101 | 622086001 |
| **Sulfonylureas (SU)** | 620006030 | 613960017 | 620009209 | 620002032 | 620002031 | 620003947 | 620003948 | 613960002 | 613960003 | 620003159 |
| | 620003160 | 610443002 | 610443003 | 621982701 | 622122201 | 622122301 | 622217701 | 610432032 | 610432033 | 610432026 |
| | 610432027 | 620001907 | 620001908 | 622462501 | 622040901 | 622041001 | | | | |
| **Sodium-glucose cotransporter type 2 inhibitors (SGLT2)** | 622306601 | 622306701 | 622341901 | 622342001 | 622335701 | 622335801 | 622336801 | 622340101 | 622360601 | 622401201 |
| | 622401301 | | | | | | | | | |
| **Dipeptidyl peptidase-4 inhibitors (DPP-4)** | 621950901 | 621951001 | 621951101 | 622277501 | 621970601 | 621970701 | 621970801 | 622288401 | 621980701 | 621986101 |
| | 621986201 | 621986001 | 622093501 | 622660601 | 622182601 | 622201701 | 622245601 | 622245701 | 622415401 | |
| **alpha-glucosidase inhibitors (aGI)** | 610406390 | 610406391 | 620002120 | 620002121 | 620005563 | 620002841 | 620002843 | 620002845 | 620002847 | 620004069 |
| | 620004070 | 620004071 | 620004072 | 620004073 | 620004074 | 620005557 | 620005558 | 620005559 | 620005560 | 620005561 |
| | 620005562 | 620005564 | 620005565 | 620005566 | 620008071 | 620008072 | 620008073 | 620008074 | 620008075 | 620008076 |
| | 620008726 | 620008728 | 621665301 | 621665401 | 621673501 | 621673601 | 621683401 | 621683501 | 621689001 | 621689101 |
| | 621689303 | 621689403 | 621690203 | 621690303 | 621690402 | 621690502 | 621690901 | 621691001 | 621691201 | 621691601 |
| | 621943301 | 621943401 | 622090001 | 622090101 | 622662501 | 622662601 | 622662701 | 622662801 | 613960081 | 613960082 |
| | 621937201 | 621937101 | 621942102 | 621942202 | 620009288 | 620009289 | 620009290 | 620009291 | 620009292 | 620009293 |
| | 620009294 | 620009295 | 620009296 | 620009297 | 621784902 | 621785002 | 621896402 | 621896502 | 620003127 | 620003128 |
| | 620003129 | 622432501 | 622426601 | 622426701 | 622560201 | 622628601 | 622560301 | 622628701 | 622560401 | 622628801 |

| | | | | | | |
|---|---|---|---|---|---|---|
| | 622664101 | 622544301 | 622664201 | 622544401 | 622664301 | 622544501 |
| **Glucagon-like peptide-1 (GLP-1) analogs** | 621974801 | 622038301 | 622038401 | 622406001 | | |
